# Effectiveness of 2025 bi-annual COVID-19 vaccination campaigns against hospitalisation in England

**DOI:** 10.64898/2026.09.14.26362989

**Authors:** Nurin Abdul Aziz, Alexander Allen, Nick Andrews

## Abstract

**Background:** In 2025, COVID-19 vaccination programmes in England targeted adults aged ≥75 years and immunosuppressed individuals, with JN.1-based vaccines offered in spring and KP.2-based vaccines offered in autumn. This study evaluated vaccine effectiveness (VE) against COVID-19 hospitalisation following these campaigns and assessed VE in immunosuppressed populations.

**Methods:** VE against COVID-19 hospitalisation in adults aged ≥75 years following the 2025 spring JN.1 and autumn KP.2 vaccination campaigns was estimated using a test-negative case-control design. Logistic regression was used to calculate VE among eligible hospitalised individuals, adjusting for covariates. Manufacturer-specific analyses were performed for the spring 2025 campaign. VE in immunosuppressed and non-immunosuppressed adults aged ≥75 years was assessed by pooling estimates from the autumn 2024 and spring and autumn 2025 campaigns using fixed-effects meta-analysis.

**Results:** Spring 2025 JN.1 vaccines conferred moderate protection, peaking at 55% (95%CI: 45-62%) within 5-9 weeks and declining to 17% (95%CI: −18-42%) from 35 weeks onwards. Autumn 2025 KP.2 VE peaked at 48% (95%CI: 34-59%) at 5-9 weeks, with follow-up needed to assess long-term protection. No significant differences were observed between manufacturers in the spring campaign. VE following autumn KP.2 vaccination was lower among individuals who received a spring JN.1 vaccine. When assessed across three campaigns, there was some evidence that VE was lower in individuals with immunosuppression than those without.

**Conclusion:** The 2025 COVID-19 vaccination programmes in England provided moderate protection against COVID-19 hospitalisation in adults aged ≥75 years, with protection waning over time. The findings support the continued benefit of seasonal vaccination in high-risk groups, including immunosuppressed individuals.

## Introduction

The epidemiology of COVID-19 has evolved since the pandemic, with the lowest levels of COVID-19 activity seen in recent periods (1,2). Despite this, COVID-19 continues to cause severe disease, and there remains a need to protect those at risk, such as older individuals and those with underlying health conditions. In 2025, SARS-CoV-2 circulation in England was initially dominated by the Omicron XEC recombinant before being replaced by JN.1.11.1 and its sub-lineages, particularly LP.8.1 and LP.8.1.1, in March 2025 (3). The emergence and subsequent dominance of XFG, a recombinant of JN.1 sub-lineage LF.7 and LP.8.12, and its sub-lineage XFG.3 occurred in June 2025, with circulation sustained to the end of the year (1,2).

Due to the waning effectiveness of previous vaccines against severe COVID-19 disease (4,5) alongside evolution of dominant SARS-CoV-2 lineages towards immune evasion, bi-annual COVID-19 vaccination programmes have been established in England to ensure continued protection for those most vulnerable to severe disease. Each programme offers updated vaccines to best match circulating lineages, based on WHO recommendations.

As with previous spring programmes in England, the spring COVID-19 campaign in 2025 targeted adults 75 years and older, residents in care homes for adults, and those aged 6 months and over who are immunosuppressed as defined by the Green Book chapter on COVID-19 (6). Vaccines offered in this campaign were the Pfizer-BioNTech and Moderna Omicron JN.1 mRNA vaccines. The programme commenced on 1 April 2025.

Eligibility for autumn 2025, which followed the same criteria as the spring 2025 campaign, was more restricted than in previous autumn programmes, which had included a broader range of individuals: in the autumn 2024 programme, adults aged 65 years and over were universally eligible, alongside those aged 6 months and over in a clinical risk group as defined in the COVID-19 Green Book chapter. The autumn 2025 campaign commenced on 1 October 2025. The products offered were Pfizer-BioNTech Omicron KP.2 mRNA vaccines for eligible individuals aged 12 years and above, and Pfizer-BioNTech Omicron LP.8.1 vaccines for eligible children aged 6 months and over.

The primary aim of this study was to estimate vaccine effectiveness (VE) and assess waning of the spring and autumn 2025 vaccines against COVID-19 hospitalisation in England using national electronic healthcare and surveillance datasets. To explore the impact of a single annual vaccination programme, we also estimated VE following the autumn 2025 campaign among individuals who had not received a spring 2025 vaccine. Finally, we compared VE in immunosuppressed and non-immunosuppressed individuals to evaluate current vaccination eligibility criteria, pooling VE estimates from the autumn 2024, spring 2025, and autumn 2025 campaigns.

## Methods

### Study design

Effectiveness of the vaccines given as part of the England spring and autumn 2025 campaigns against hospitalisation was calculated using the test-negative case-control (TNCC) study design where positive SARS-CoV-2 polymerase chain reaction (PCR) tests from hospitalised individuals are cases and negative PCR tests from hospitalised individuals are controls, as previously described. The study period was from 1 April 2025 to 5 April 2026 for the spring campaign, and 1 October 2025 to 5 April 2026 for the autumn campaign, with data extracted on 8 May 2026.

Tests from the autumn 2024, spring 2025, and autumn 2025 vaccination campaigns were included in the analysis comparing VE between immunosuppressed and non-immunosuppressed individuals. The study period for autumn 2024 was from 3 October 2024 to 26 October 2025, with the other campaign study periods as above.

### Data sources

#### SARS-CoV-2 testing data

Positive and negative SARS-CoV-2 PCR tests conducted in hospital settings in England were identified through the Unified Sample Dataset (USD), a national repository for SARS-CoV-2 data, as previously described (5,7). Testing data is enhanced with linkage to the NHS spine to add patient identifiers, including NHS number.

Initially, tests were deduplicated such that individuals contributed up to one negative test and one positive test per day. Where multiple tests occurred on the same day, positive results were prioritised over negative. From the deduplicated tests, any test, positive or negative, within 90 days after a positive test was excluded, as these likely represent the same episode. Negative tests were excluded if within 21 days before a positive test. Eligible individuals could contribute a maximum of one randomly selected negative test per vaccination campaign period; hence, for assessment of spring 2025 VE, an individual could contribute up to two negative tests: one during the spring/summer period and one during the autumn/winter period. Random selection of negatives occurred only among hospital-linked tests. Individuals could contribute more than one positive, but the above criteria mean they must be at least 90 days apart. Test exclusion criteria and rationale have been described previously (8,9).

#### COVID-19 vaccination data

To obtain data on individual vaccination history, testing data was linked to the UKHSA Immunisation Information System (IIS), a national vaccine register containing demographic information and COVID-19 and influenza vaccination records for individuals living in England who are registered with a general practitioner (10). Vaccine data included dates of vaccination, vaccine type, and manufacturer.

Doses distributed through the campaigns were classified based on vaccine products derived from vaccine batch number and timing of vaccination. Tests were excluded if: the individual received a dose from a manufacturer other than those primarily offered; the individual had two or more doses within the campaign being analysed; or if there was less than 12 weeks between the dose received within the campaign and their previous dose. For analyses focusing on waning of the spring 2025 campaign, individuals who received an autumn 2025 dose prior to the test date were removed.

#### Demographic and clinical risk data

Linkage to the IIS provided demographic information, including sex, age, ethnicity, and residential address. Postcodes were used to determine index of multiple deprivation (IMD), an area-based measure of relative deprivation.

Clinical risk status at the start of each vaccination campaign was obtained from the NHS Cohorting as a Service (CaaS) platform, which identifies individuals eligible for vaccination on the basis of clinical risk, including immunosuppression, as defined in the COVID-19 Green Book (6). While data on a wider range of clinical risk groups were available for campaigns up to spring 2025, only immunosuppression indicators were available for the autumn 2025 campaign because eligibility in both campaigns was limited to older age groups and individuals who were immunosuppressed.

For analyses comparing VE between immunosuppressed and non-immunosuppressed populations, non-immunosuppressed individuals were those with either no identified clinical risk or those in clinical risk groups other than immunosuppression.

#### COVID-19 hospitalisation data

Data on hospital admissions was obtained from the Secondary Uses Services (SUS) data set, a national administrative data set capturing information on NHS hospital and ICU admissions in England to inform management and planning of NHS services. This data contained dates of admission and discharge, ICD-10 diagnosis codes, procedure codes, and specialty and treatment function codes.

Testing data was linked to hospital admission records using NHS number and date of birth. Completeness of SUS data follows a lag of a few weeks; therefore, testing data was restricted to approximately 3 weeks prior to the date of data extraction and linkage.

Hospital admissions were defined as any admission 2 days before up to 1 day after a COVID-19 test, where the admission record was associated with an ICD-10 coded acute respiratory illness diagnosis in the primary diagnosis field, and where the admission length was at least 2 days, calculated as the difference between discharge date and admission date. Only the first admission was retained where multiple admissions occurred within 21 days of a test.

### Statistical analysis

#### Effectiveness of spring and autumn 2025 vaccines

VE estimates were calculated separately for the spring and autumn 2025 campaigns. Multivariable logistic regression was used to calculate odds ratios (OR), with time interval since vaccination in the respective campaign as the exposure, and SARS-CoV-2 test result as the outcome. VE was calculated as 1-OR with 95% confidence intervals (CIs). Statistical significance when comparing VE estimates was concluded where 95% CIs did not overlap. This approach is conservative (approximately equivalent to a significance level of 0.5-1%) but allows comparison of multiple estimates.

Incremental VE of the spring 2025 JN.1 vaccine was estimated for those aged 75 years and older at the following intervals since vaccination: 0–8 days, 9–13 days, 2–4 weeks, 5–9 weeks, 10–14 weeks, 15–19 weeks, 20–24 weeks, 25–29 weeks, 30–34 weeks, and ≥35 weeks. Separate analyses were conducted to assess VE stratified by manufacturer.

Similarly, incremental VE of the autumn 2025 KP.2 vaccine was estimated for those aged 75 years and older at the same intervals as the spring campaign up to ≥20 weeks. VE was not stratified by manufacturer as only one manufacturer was represented in this campaign.

Analyses included adjustment for: 5-year age group at the start of the vaccination campaign as a categorical variable; week of SARS-CoV-2 test as a categorical variable; sex; NHS region; IMD quintile; ethnicity; and influenza vaccination in the most recent season. Adjustment for clinical risk group was also included: for the spring campaign, clinical risk group status was coded as no evidence of clinical risk, clinical risk other than immunosuppression, and immunosuppressed; for the autumn campaign, clinical risk group status was coded as no immunosuppression risk, and immunosuppressed.

#### VE by previous campaign vaccination status

To investigate the impact of a twice-yearly vaccination programme compared with a single annual campaign, we examined whether autumn 2025 VE, calculated as described above, differed between hospitalised individuals who had and had not received a spring 2025 vaccine. Time since receipt of an autumn 2025 dose was stratified into wider intervals of 0–13 days, 2–19 weeks, and ≥20 weeks to increase statistical power. Likelihood ratio tests were conducted to assess whether receipt of a spring 2025 vaccine dose (yes/no) significantly modified the effectiveness estimates over time.

#### VE by immunosuppression status

VE estimates were calculated, as described above, for the autumn 2024, spring 2025, and autumn 2025 campaigns for hospitalised individuals aged 75 and over who were flagged as immunosuppressed, and separately those who did not have an immunosuppression flag. Time since receipt of the respective campaign dose was stratified into wider intervals of 0–13 days, 2–19 weeks, and ≥20 weeks to increase statistical power.

Season-specific estimates were pooled using fixed-effects meta-analysis to account for limited sample sizes among immunosuppressed individuals. Season-specific log ORs from the logistic regression models were combined, and the resulting pooled estimates were used to calculate VE at each time interval for the immunosuppressed and non-immunosuppressed groups.

## Results

There were 41,928 SARS-CoV-2 tests eligible for the spring 2025 VE analysis during the study period, of which 4,503 were cases and 37,426 were controls. For the autumn 2025 study, 35,674 tests were eligible for the analysis, of which 1,994 were cases and 33,680 were controls. Full descriptive characteristics of the study population (Supplementary Tables 1 and 2) and a distribution of cases and controls over time (Supplementary Figs. 1-5) for both the spring and autumn campaigns can be found in the supplementary document.

### Effectiveness of the spring 2025 JN.1 vaccines

Protection against hospitalisation provided by the JN.1 vaccine given in the spring 2025 campaign in adults aged ≥75 years was moderate (Figure 1). VE peaked at 55% (95%CI: 45-62%) 5-9 weeks after vaccination and decreased to 33% (95%CI: 22-43%) by 10-14 weeks. By 35 weeks and onwards, there is some evidence of waning to 17% (95%CI: −18-42%).

**Figure 1.**
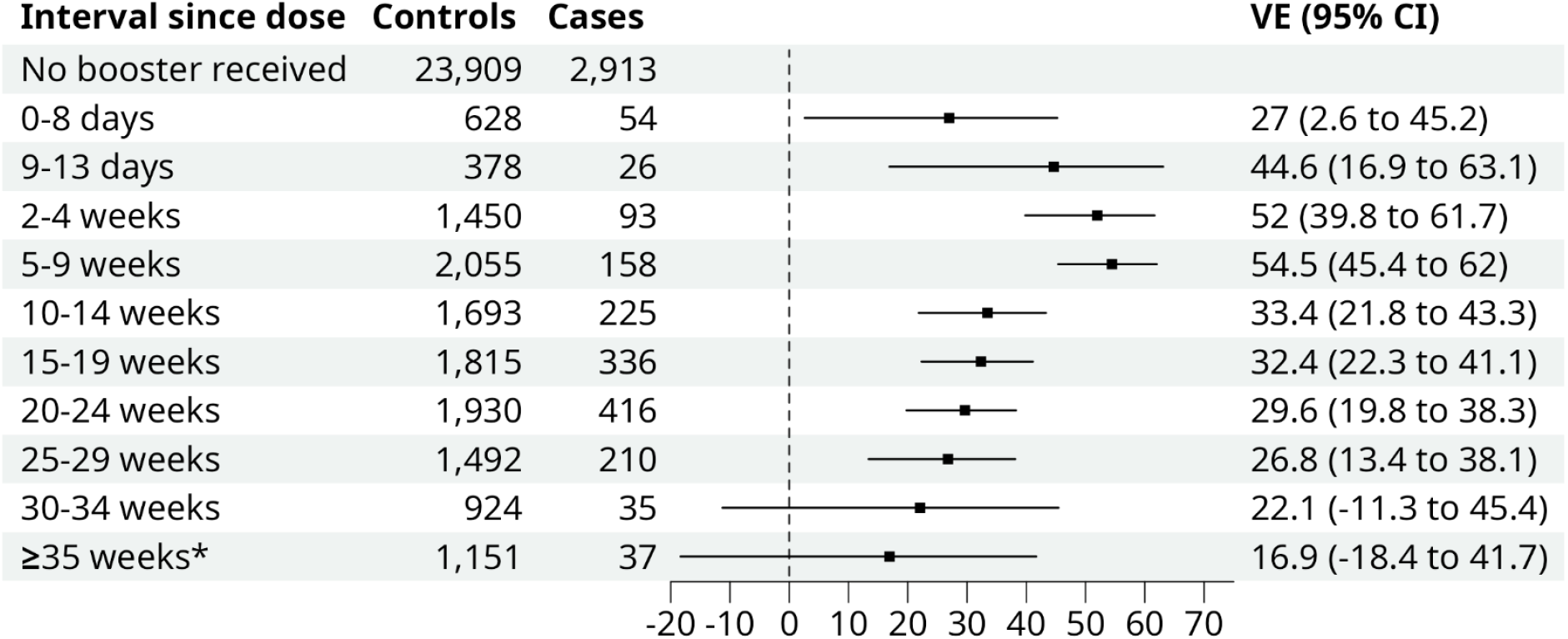
Vaccine effectiveness (VE) of COVID-19 spring 2025 JN.1 vaccines against hospitalisation in adults aged ≥75 years, stratified by time since vaccination. Error bars represent 95% confidence intervals (CI). *Median for time since dose 35 weeks or greater: 39 weeks, IQR: 36-42 weeks.

Among eligible tests from recipients of the spring campaign vaccine, 79% were from individuals who received Moderna JN.1 and 21% from those who received Pfizer-BioNTech JN.1. The peak Moderna JN.1 VE estimate (54%, 95%CI: 44–63%) was marginally higher than the peak Pfizer JN.1 VE estimate (47%, 95%CI: 24–63%), although the overlapping CIs indicate no significant differences (Figure 2). By ≥35 weeks, VE for Moderna JN.1 fell to 17% (95%CI: −21-43%). Pfizer JN.1 VE fell to 35% (95%CI: −76-67%) in the same interval. Estimates from 25 weeks onwards had wide CIs, limiting interpretation of any waning. Despite slight differences in point estimates, CIs for both vaccines overlapped at all time points, indicating limited evidence of differing VE between the two vaccines.

**Figure 2.**
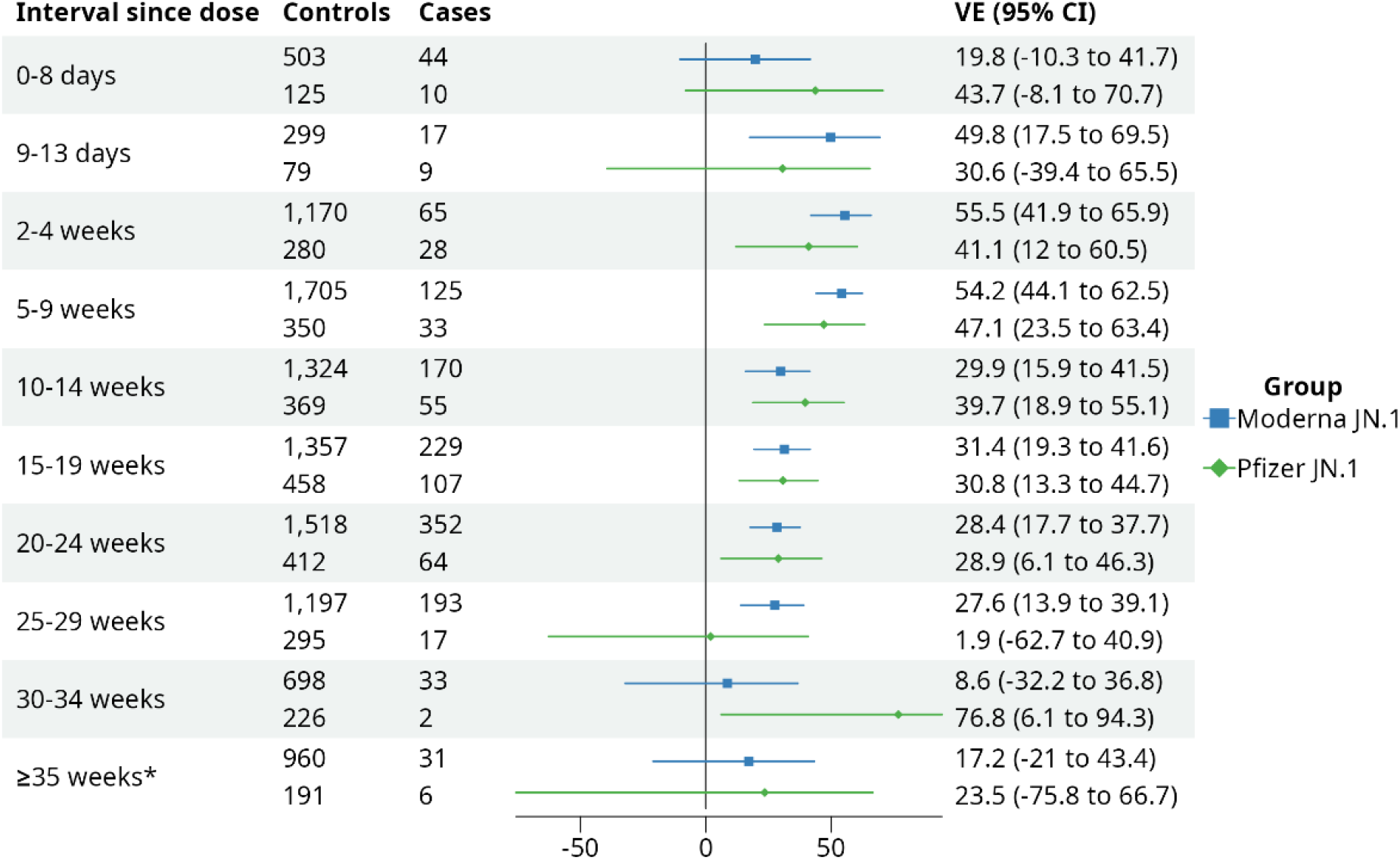
Vaccine effectiveness (VE) of COVID-19 spring 2025 JN.1 vaccines against hospitalisation in adults aged ≥75 years, stratified by manufacturer (Moderna and Pfizer) and time since vaccination. Error bars represent 95% confidence intervals (CI). *Median for time since dose 35 weeks or greater: Moderna – 39 weeks, IQR: 37-42 weeks; Pfizer – 38 weeks, IQR: 36-40 weeks.

### Effectiveness of the autumn 2025 KP.2 vaccines

The autumn KP.2 vaccine also showed moderate protection against hospitalisation (Figure 3) in adults aged ≥75 years, peaking at 5-9 weeks after vaccination at 48% (95%CI: 34-59%). There was some evidence of waning at ≥20 weeks after vaccination, with VE decreasing to 35% (95%CI: −1-59%); however, the wide CIs at longer intervals post-vaccination indicate additional follow-up will be needed to better characterise the extent of waning.

**Figure 3.**
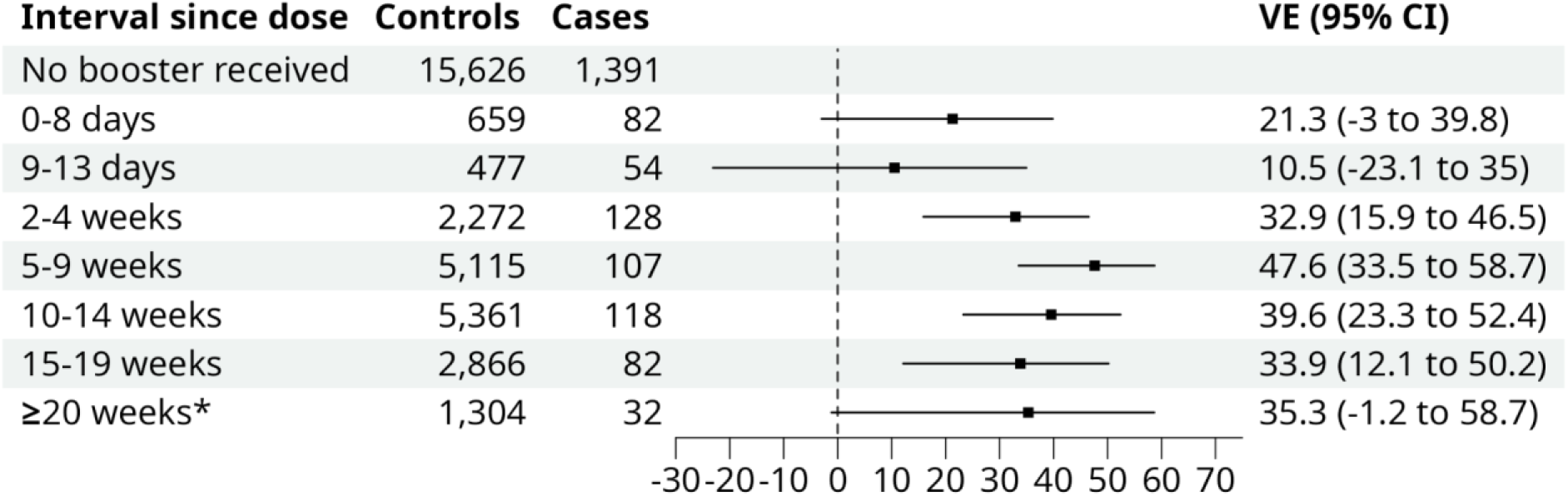
Vaccine effectiveness (VE) of COVID-19 autumn 2025 KP.2 vaccines against hospitalisation in adults aged ≥75 years, stratified by time since vaccination. Error bars represent 95% confidence intervals (CI). *Median for time since dose 20 weeks or greater: 21 weeks, IQR: 20-23 weeks.

Effectiveness of the autumn KP.2 vaccine against hospitalisation at 2-19 weeks after vaccination is moderately higher in individuals aged ≥75 years who have not previously received a spring JN.1 vaccine (42%, 95%CI: 27-54%) compared to those who have received the JN.1 vaccine (31%, 95%CI: 17-43%) (Figure 4). However, overlapping CIs indicate limited evidence of a difference in peak VE between the two groups. Additionally, wide CIs and small sample sizes at 20 weeks or more after vaccination for both groups limit comparisons of VE. Despite this, there was a significant interaction effect between receipt of a spring dose and time since vaccination (p = 0.005), suggesting VE varies differently over time between the two groups.

**Figure 4.**
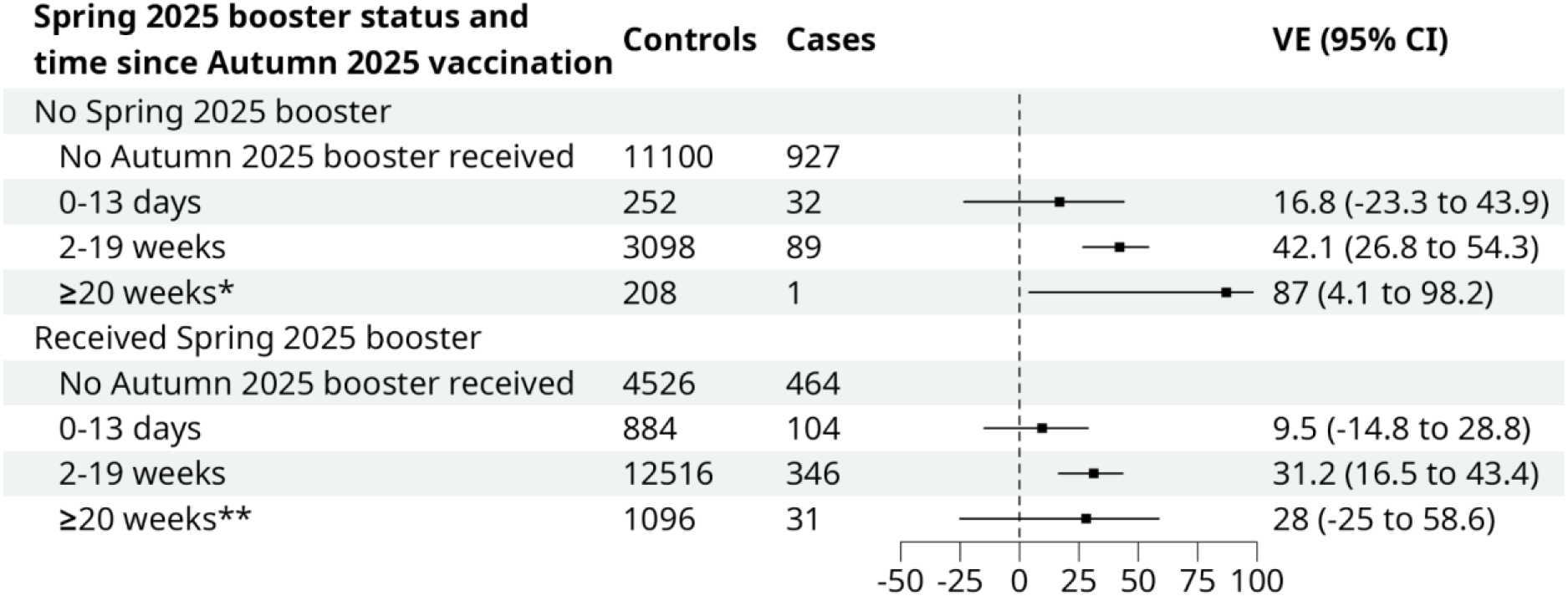
Vaccine effectiveness (VE) of COVID-19 autumn 2025 KP.2 vaccines against hospitalisation in adults aged ≥75 years, stratified by time since vaccination and receipt of spring 2025 vaccine. Error bars represent 95% confidence intervals (CI). *Median for time since dose 20 weeks or greater for individuals without a spring 2025 dose: 22 weeks, IQR: 21-23 weeks. **Median for time since dose 20 weeks or greater for individuals with a spring 2025 dose: 21 weeks, IQR: 20-23 weeks.

### Effectiveness by immunosuppression status

Pooled VE estimates of the autumn 2024, spring 2025, and autumn 2025 campaigns across all intervals since vaccination were lower for immunosuppressed individuals aged ≥75 years compared to non-immunosuppressed (Figure 5). Distribution of cases and controls and total counts of tests used in the analysis can be found in Supplementary Figure 6 and Supplementary Table 3. Peak VE was seen at 2-19 weeks for both groups at 33% (95%CI: 21-42%) for immunosuppressed and 41% (95%CI: 36-44%) for non-immunosuppressed. There was also lower VE in the immunosuppressed group at 20-35 weeks. The combined VE at 2-9 weeks and 20-35+ weeks is 35% (95%CI: 32-38%) for the non-immunosuppressed, and 25% for the immunosuppressed (95%CI: 18-35%). Although there is a 10% VE difference, the CIs overlapping to this extent indicate this is not significant.

**Figure 5.**
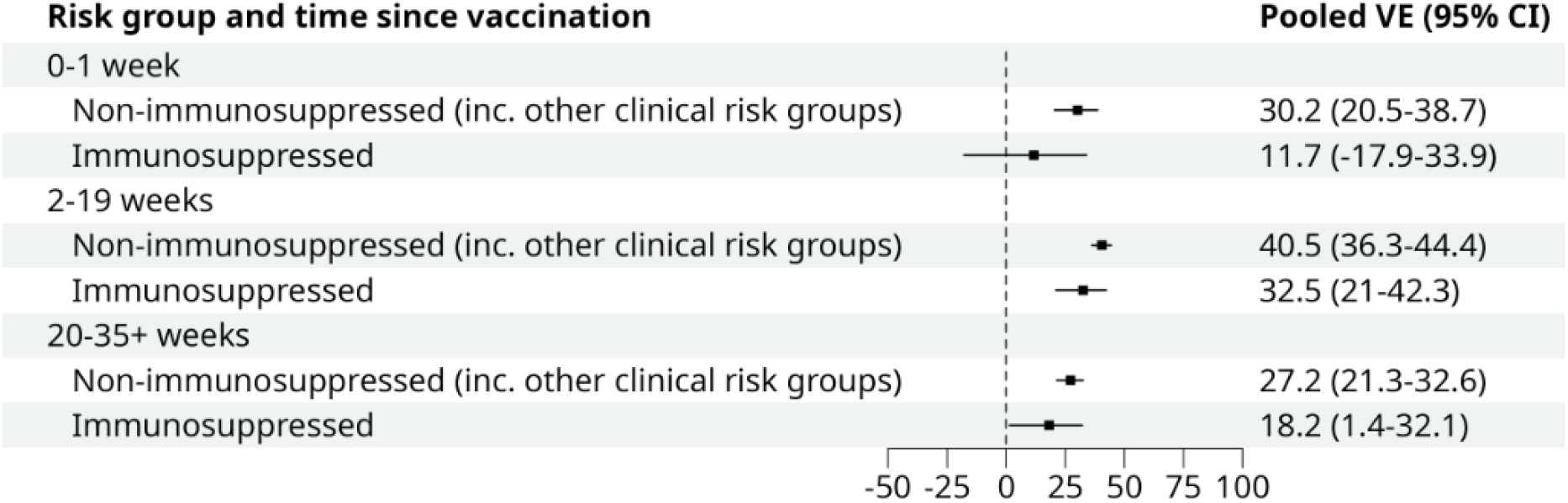
Pooled vaccine effectiveness (VE) estimates from a fixed-effects model aggregating VE estimates from autumn 2024, spring 2025, and autumn 2025 campaigns in adults aged ≥75 years, stratified by time since vaccination and immunosuppression status.

## Discussion

This study evaluated the effectiveness of COVID-19 vaccines given during the spring and autumn 2025 campaigns against hospitalisation among older adults in England using a TNCC design. We find the spring JN.1 vaccine and the autumn KP.2 vaccine conferred moderate protection against hospitalisation in addition to any remaining protection from previous vaccinations, with VE peaking at 55% and 48% respectively.

Rollout of the spring JN.1 vaccine occurred during an LP.8.1 and XFG-dominant season. JN.1 vaccines were seen to boost neutralisation against LP.8.1 in virological characterisation and immunogenicity trials, though at lower levels than against the parent JN.1 lineage, which may contribute to the moderate protection (11). Similar estimates for the JN.1 vaccine against JN.1 sub-lineages were seen in other countries. A TNCC study based in Japan showed comparable peak VE against hospitalisation of 55% (95%CI: −27-84%) 7-60 days after vaccination, though with wide CIs at this interval (12). A cohort study in the United States showed peak VE against hospitalisation of 57% (95%CI: 43-68%) at 4 weeks; similar effectiveness was seen against circulating sub-variants, including the emerging LP.8.1 (13). In an EU/EEA network cohort study, peak VE of the JN.1 vaccine against hospitalisation was 60% (95%CI: 48-70%) against a backdrop of KP.3.1.1 and XEC circulation (14).

Peak protection from the spring JN.1 vaccine was seen in the first 2 months after vaccination, with significant protection sustained until at least week 30. This is consistent with a previous analysis of the autumn 2024 JN.1 campaign, which also demonstrated sustained effectiveness against hospitalisation, in contrast to the more rapid waning seen in previous campaigns (1). This potentially suggests duration of protection may have increased in recent campaigns.

In previous seasons, there has been increased circulation of novel SARS-CoV-2 variants with a marked increase in immune escape capability, in addition to large antigenic shifts in circulating variants. In comparison, the expansion of LP.8.1 was better explained through increased receptor-binding affinity and not associated with increased immune escape, which may contribute to the sustained protection provided by the JN.1 vaccine (15). XFG conversely showed reduced antibody sensitivity compared to LP.8.1.1, though not as much as previous emergent variants (16). The autumn 2024 campaign was rolled out during a period when XEC was the dominant variant, followed by the expansion and subsequent dominance of LP.8.1 (17). Again, the sustained protection observed may partly reflect the relatively limited increase in immune escape associated with LP.8.1 and the similarity of its immune resistance profile to that of its predecessor, XEC (18). Furthermore, increased opportunities for infection over time may enhance hybrid immunity, potentially improving antibody response durability and extending vaccine protection in later campaigns (19,20).

The autumn 2025 campaign occurred during a period of XFG dominance; moderate levels of protection may have been caused by reduced neutralisation of XFG by the KP.2 vaccine (21). While rollout of the KP.2 vaccine in other countries occurred earlier than in England and therefore covered a period of different circulating variants, including KP.3.1.1 and XEC, similar estimates of effectiveness against hospitalisation were seen in two TNCC studies from the United States: peak VE of 49% (95%CI: 30-63%) and 40% (95%CI: 27-51%) were presented by the Kaiser Permanente Southern California group (22) and the IVY Network respectively (23). Further follow-up is required to understand the duration of protection for the autumn KP.2 vaccine.

The peak effectiveness estimates for the autumn 2025 KP.2 vaccine were slightly lower than the spring 2025 JN.1 vaccine, though overlapping CIs do not indicate a statistically significant difference. One possible explanation is residual protection from the spring 2025 vaccine: as the eligibility criteria for the two campaigns were the same, many individuals vaccinated in autumn 2025 are likely to have received a spring 2025 vaccine previously, which may have conferred modest residual protection. This is supported by the analysis stratifying prior vaccination status, in which peak autumn 2025 VE was marginally lower among individuals who had received a spring 2025 vaccine, at 32%, than among those who had not, at 42%. However, the overlapping CIs between the estimates indicate uncertainty around these estimates, and additional follow-up is needed to better understand the contribution of residual protection. This evidence will help inform the impact of biannual vaccination campaigns and the additional protection provided by a second vaccination within the same year, accounting for potential changes in duration of waning.

Furthermore, this campaign occurred shortly after the only peak of COVID-19 activity in 2025 (1,2), which may have increased overall population-level immunity derived from infections, resulting in lower incremental protection from the vaccine; conversely, the wave prior to the spring 2025 campaign was in October 2024, which may have resulted in lower population-level immunity at the point of the campaign.

There was some evidence that pooled effectiveness of vaccines rolled out in the autumn 2025, spring 2025, and autumn 2024 seasons appears to be lower in the immunosuppressed population, with peak VE in the immunosuppressed at 33% against 41% in the non-immunosuppressed. This effect has been corroborated in previous literature, where a reduced immune response to COVID-19 vaccines was observed in immunosuppressed individuals (24,25); nevertheless, this still represents some evidence of protection for this population. Additionally, overlapping CIs between the two populations limits interpretation of the differences in VE, and further data from the immunosuppressed population may be required.

The TNCC study design has been widely used to assess VE against COVID-19 at the population-level. Its key strength lies in addressing unmeasured confounders related to health-seeking behaviours, healthcare access, and exposure to infection. This study provides the first evidence of the effectiveness of the JN.1 vaccine against COVID-19 hospitalisation during periods of predominant LP.8.1 and XFG circulation, and of the KP.2 vaccine during predominant XFG circulation. It also provides updated estimates of VE in immunosuppressed populations using data extending to 2026.

This study also has several limitations. As this is an observational study, there may remain unmeasured or residual confounding. National-level healthcare datasets may also result in variable data quality: incomplete linkage may result in missing data, and errors in coding within hospital data may result in incorrect inclusion of non-respiratory admissions within the study.

Low COVID-19 activity throughout 2025 limits sample sizes and subsequently reduces statistical power. Due to the scale-down of genomic surveillance, it is not fully possible to untangle whether changes in VE over time is due to waning vaccine effects or changes in immune evasion properties associated with new variants. Additionally, the lack of genomic data limits the ability to assess variant-specific VE, which is important for evaluating variant-specific vaccine updates. Currently, VE is evaluated against hospitalisations occurring during periods when specific variants are dominant. More comprehensive genomic data would enable direct assessment of protection against individual variants and provide stronger evidence on whether updating vaccine composition improves protection against COVID-19 hospitalisations.

Another limitation can be seen in the exclusion criteria for the spring 2025 analysis, where we remove individuals who received an autumn 2025 dose prior to their test date. The population may hence change midway through the study period, wherein those who have received a spring vaccine but not an autumn vaccine may have different healthcare seeking behaviours compared to those who sought and received both vaccines. The long-term waning assessment could be confounded by this changing population.

In conclusion, this study provides evidence of moderate protection against hospitalisations conferred by the JN.1 vaccine and the KP.2 vaccine distributed in England during the COVID-19 spring 2025 and autumn 2025 campaigns respectively within older adults eligible for the vaccines, with protection sustained for at least 20 weeks after vaccination.

## Statements

### Data availability

Vaccine effectiveness work is carried out under Regulation 3 of The Health Service (Control of Patient Information; Secretary of State for Health, 2002), which includes the use of patient identification information without individual patient consent as part of the UKHSA legal requirement for public health surveillance and monitoring of vaccines. Authors cannot make the underlying dataset publicly available for ethical and legal reasons. However, data used for this analysis are included as aggregated data in the manuscript tables and appendix. Applications for relevant anonymised data should be submitted to the UKHSA Office for Data Release: https://www.gov.uk/government/publications/accessing-ukhsa-protected-data.

### Ethical statement

The study protocol was subject to an internal review by the UK Health Security Agency Research Ethics and Governance Group and was found to be fully compliant with all regulatory requirements. As no regulatory issues were identified, and ethical review is not a requirement for this type of work, it was decided that a full ethical review would not be necessary.

UKHSA has legal permission, provided by Regulation 3 of The Health Service (Control of Patient Information) Regulations 2002, to process patient confidential information for national surveillance of communicable diseases and as such, individual patient consent is not required to access records.

