## Supplementary Information for "Effectiveness of 2025 bi-annual COVID-19 vaccination campaigns against hospitalisation in England"

Supplementary Table 1. Descriptive characteristics of cases and controls included in the analysis of the effectiveness of the JN.1 vaccine given in spring 2025 against hospitalisation amongst individuals aged 75 years and older in England.

|  | **Controls** | | **Cases** | | **Overall** | |
| --- | --- | --- | --- | --- | --- | --- |
|  | n | % | n | % | n | % |
| **Interval since booster dose** | | | | | | |
| No booster | 23,909 | 63.89 | 2,913 | 64.69 | 26,822 | 63.97 |
| 0-8 days | 628 | 1.68 | 54 | 1.20 | 682 | 1.63 |
| 9-13 days | 378 | 1.01 | 26 | 0.58 | 404 | 0.96 |
| 2-4 weeks | 1,450 | 3.87 | 93 | 2.07 | 1,543 | 3.68 |
| 5-9 weeks | 2,055 | 5.49 | 158 | 3.51 | 2,213 | 5.28 |
| 10-14 weeks | 1,693 | 4.52 | 225 | 5.00 | 1,918 | 4.57 |
| 15-19 weeks | 1,815 | 4.85 | 336 | 7.46 | 2,151 | 5.13 |
| 20-24 weeks | 1,930 | 5.16 | 416 | 9.24 | 2,346 | 5.60 |
| 25-29 weeks | 1,492 | 3.99 | 210 | 4.66 | 1,702 | 4.06 |
| 30-34 weeks | 924 | 2.47 | 35 | 0.78 | 959 | 2.29 |
| ≥35 weeks (Median: 39 weeks, IQR: 36-42 weeks) | 1,151 | 3.08 | 37 | 0.82 | 1,188 | 2.83 |
| **Age group** | | | | | | |
| 75-79 | 10,574 | 28.25 | 1,163 | 25.83 | 11,737 | 27.99 |
| 80-84 | 10,469 | 27.97 | 1,280 | 28.43 | 11,749 | 28.02 |
| 85-89 | 9,326 | 24.92 | 1,137 | 25.25 | 10,463 | 24.95 |
| ≥90 | 7,056 | 18.85 | 923 | 20.50 | 7,979 | 19.03 |
| **Sex** | | | | | | |
| Female | 19,779 | 52.85 | 2,224 | 49.39 | 22,003 | 52.48 |
| Male | 16,993 | 45.41 | 2,266 | 50.32 | 19,259 | 45.93 |
| Unknown | 653 | 1.74 | 13 | 0.29 | 666 | 1.59 |
| **Ethnic group** | | | | | | |
| African (Black or Black British) | 141 | 0.38 | 17 | 0.38 | 158 | 0.38 |
| Any other Asian background | 338 | 0.90 | 37 | 0.82 | 375 | 0.89 |
| Any other Black background | 91 | 0.24 | 19 | 0.42 | 110 | 0.26 |
| Any other ethnic group | 343 | 0.92 | 55 | 1.22 | 398 | 0.95 |
| Any other Mixed background | 81 | 0.22 | 12 | 0.27 | 93 | 0.22 |
| Any other White background | 1,768 | 4.72 | 219 | 4.86 | 1,987 | 4.74 |
| Bangladeshi (Asian or Asian British) | 169 | 0.45 | 15 | 0.33 | 184 | 0.44 |
| British (White) | 30,479 | 81.44 | 3,701 | 82.19 | 34,180 | 81.52 |
| Caribbean (Black or Black British) | 327 | 0.87 | 41 | 0.91 | 368 | 0.88 |
| Chinese (other ethnic group) | 67 | 0.18 | 3 | 0.07 | 70 | 0.17 |
| Indian (Asian or Asian British) | 832 | 2.22 | 77 | 1.71 | 909 | 2.17 |
| Irish (White) | 513 | 1.37 | 58 | 1.29 | 571 | 1.36 |
| Pakistani (Asian or Asian British) | 496 | 1.33 | 42 | 0.93 | 538 | 1.28 |
| White and Asian (Mixed) | 23 | 0.06 | 5 | 0.11 | 28 | 0.07 |
| White and Black African (Mixed) | 23 | 0.06 | 3 | 0.07 | 26 | 0.06 |
| White and Black Caribbean (Mixed) | 46 | 0.12 | 4 | 0.09 | 50 | 0.12 |
| Unknown | 1,688 | 4.51 | 195 | 4.33 | 1,883 | 4.49 |
| **IMD quintile** | | | | | | |
| 1 | 7,933 | 21.20 | 879 | 19.52 | 8,812 | 21.02 |
| 2 | 7,556 | 20.19 | 956 | 21.23 | 8,512 | 20.30 |
| 3 | 7,473 | 19.97 | 938 | 20.83 | 8,411 | 20.06 |
| 4 | 7,465 | 19.95 | 878 | 19.50 | 8,343 | 19.90 |
| 5 | 6,815 | 18.21 | 831 | 18.45 | 7,646 | 18.24 |
| Unknown | 183 | 0.49 | 21 | 0.47 | 204 | 0.49 |
| **Region** | | | | | | |
| East of England | 3,470 | 9.27 | 396 | 8.79 | 3,866 | 9.22 |
| London | 4,969 | 13.28 | 611 | 13.57 | 5,580 | 13.31 |
| Midlands | 8,730 | 23.33 | 1,031 | 22.90 | 9,761 | 23.28 |
| North East and Yorkshire | 6,982 | 18.66 | 918 | 20.39 | 7,900 | 18.84 |
| North West | 5,631 | 15.05 | 538 | 11.95 | 6,169 | 14.71 |
| South East | 3,857 | 10.31 | 492 | 10.93 | 4,349 | 10.37 |
| South West | 3,782 | 10.11 | 517 | 11.48 | 4,299 | 10.25 |
| Unknown | 4 | 0.01 | 0 | 0.00 | 4 | 0.01 |
| **Clinical risk status** | | | | | | |
| None | 3,345 | 8.94 | 358 | 7.95 | 3,703 | 8.83 |
| Clinical risk group, not immunosuppressed | 28,911 | 77.25 | 3,492 | 77.55 | 32,403 | 77.28 |
| Immunosuppressed | 5,169 | 13.81 | 653 | 14.50 | 5,822 | 13.89 |
| **Flu vaccination status** | | | | | | |
| None | 10,590 | 28.30 | 1,237 | 27.47 | 11,827 | 28.21 |
| Influenza vaccine received | 26,835 | 71.70 | 3,266 | 72.53 | 30,101 | 71.79 |

Supplementary Table 2. Descriptive characteristics of cases and controls included in the analysis of the effectiveness of the KP.2 vaccine given in autumn 2025 against hospitalisation amongst individuals aged 75 years and older in England.

|  | **Controls** | | **Cases** | | | **Overall** | | |
| --- | --- | --- | --- | --- | --- | --- | --- | --- |
|  | n | % | | n | % | | n | % |
| **Interval since booster dose** | | | | | | | | |
| No booster | 15,626 | 46.40 | | 1,391 | 69.76 | | 17,017 | 47.70 |
| 0-2 days | 143 | 0.42 | | 21 | 1.05 | | 164 | 0.46 |
| 3-8 days | 516 | 1.53 | | 61 | 3.06 | | 577 | 1.62 |
| 9-13 days | 477 | 1.42 | | 54 | 2.71 | | 531 | 1.49 |
| 2-4 weeks | 2,272 | 6.75 | | 128 | 6.42 | | 2,400 | 6.73 |
| 5-9 weeks | 5,115 | 15.19 | | 107 | 5.37 | | 5,222 | 14.64 |
| 10-14 weeks | 5,361 | 15.92 | | 118 | 5.92 | | 5,479 | 15.36 |
| 15-19 weeks | 2,866 | 8.51 | | 82 | 4.11 | | 2,948 | 8.26 |
| ≥20 weeks (Median: 21 weeks, IQR: 20-23 weeks) | 1,304 | 3.87 | | 32 | 1.60 | | 1,336 | 3.75 |
| **Age group** | | | | | | | | |
| 75-79 | 9,455 | 28.07 | | 543 | 27.23 | | 9,998 | 28.03 |
| 80-84 | 9,691 | 28.77 | | 575 | 28.84 | | 10,266 | 28.78 |
| 85-89 | 8,672 | 25.75 | | 500 | 25.08 | | 9,172 | 25.71 |
| ≥90 | 5,862 | 17.40 | | 376 | 18.86 | | 6,238 | 17.49 |
| **Sex** | | | | | | | | |
| Female | 17,856 | 53.02 | | 1,004 | 50.35 | | 18,860 | 52.87 |
| Male | 15,330 | 45.52 | | 989 | 49.60 | | 16,319 | 45.74 |
| Unknown | 494 | 1.47 | | 1 | 0.05 | | 495 | 1.39 |
| **Ethnic group** | | | | | | | | |
| African (Black or Black British) | 89 | 0.26 | | 5 | 0.25 | | 94 | 0.26 |
| Any other Asian background | 212 | 0.63 | | 7 | 0.35 | | 219 | 0.61 |
| Any other Black background | 57 | 0.17 | | 5 | 0.25 | | 62 | 0.17 |
| Any other ethnic group | 235 | 0.70 | | 14 | 0.70 | | 249 | 0.70 |
| Any other Mixed background | 69 | 0.20 | | 5 | 0.25 | | 74 | 0.21 |
| Any other White background | 1,435 | 4.26 | | 83 | 4.16 | | 1,518 | 4.26 |
| Bangladeshi (Asian or Asian British) | 105 | 0.31 | | 4 | 0.20 | | 109 | 0.31 |
| British (White) | 28,332 | 84.12 | | 1,716 | 86.06 | | 30,048 | 84.23 |
| Caribbean (Black or Black British) | 242 | 0.72 | | 16 | 0.80 | | 258 | 0.72 |
| Chinese (other ethnic group) | 48 | 0.14 | | 3 | 0.15 | | 51 | 0.14 |
| Indian (Asian or Asian British) | 597 | 1.77 | | 25 | 1.25 | | 622 | 1.74 |
| Irish (White) | 451 | 1.34 | | 17 | 0.85 | | 468 | 1.31 |
| Pakistani (Asian or Asian British) | 326 | 0.97 | | 16 | 0.80 | | 342 | 0.96 |
| White and Asian (Mixed) | 22 | 0.07 | | 1 | 0.05 | | 23 | 0.06 |
| White and Black African (Mixed) | 18 | 0.05 | | 1 | 0.05 | | 19 | 0.05 |
| White and Black Caribbean (Mixed) | 37 | 0.11 | | 1 | 0.05 | | 38 | 0.11 |
| Unknown | 1,405 | 4.17 | | 75 | 3.76 | | 1,480 | 4.15 |
| **IMD quintile** | | | | | | | | |
| 1 | 6,293 | 18.68 | | 353 | 17.70 | | 6,646 | 18.63 |
| 2 | 6,540 | 19.42 | | 385 | 19.31 | | 6,925 | 19.41 |
| 3 | 6,796 | 20.18 | | 431 | 21.61 | | 7,227 | 20.26 |
| 4 | 7,083 | 21.03 | | 415 | 20.81 | | 7,498 | 21.02 |
| 5 | 6,808 | 20.21 | | 398 | 19.96 | | 7,206 | 20.20 |
| Unknown | 160 | 0.48 | | 12 | 0.60 | | 172 | 0.48 |
| **Region** | | | | | | | | |
| East of England | 3,137 | 9.31 | | 178 | 8.93 | | 3,315 | 9.29 |
| London | 3,614 | 10.73 | | 162 | 8.12 | | 3,776 | 10.58 |
| Midlands | 7,559 | 22.44 | | 436 | 21.87 | | 7,995 | 22.41 |
| North East and Yorkshire | 6,471 | 19.21 | | 441 | 22.12 | | 6,912 | 19.38 |
| North West | 4,764 | 14.14 | | 235 | 11.79 | | 4,999 | 14.01 |
| South East | 3,855 | 11.45 | | 248 | 12.44 | | 4,103 | 11.50 |
| South West | 4,280 | 12.71 | | 294 | 14.74 | | 4,574 | 12.82 |
| **Clinical risk status** | | | | | | | | |
| Non-immunosuppressed | 28,019 | 83.19 | | 1,621 | 81.29 | | 29,640 | 83.09 |
| Immunosuppressed | 5,661 | 16.81 | | 373 | 18.71 | | 6,034 | 16.91 |
| **Flu vaccination status** | | | | | | | | |
| None | 11,179 | 33.19 | | 1,171 | 58.73 | | 12,350 | 34.62 |
| Influenza vaccine received | 22,501 | 66.81 | | 823 | 41.27 | | 23,324 | 65.38 |

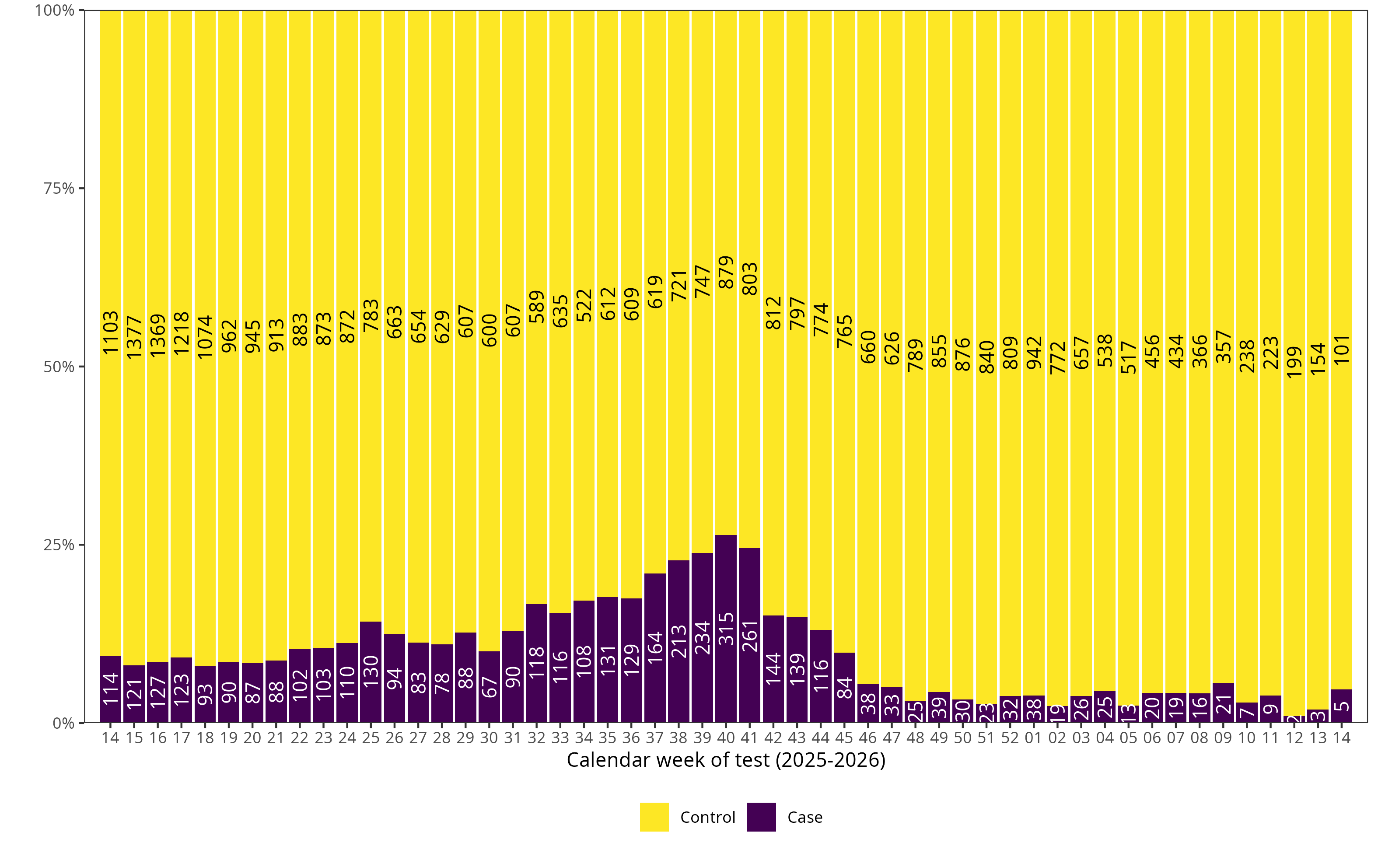

Supplementary Figure 1. Distribution of cases and controls by week of test for tests included in the spring 2025 vaccine effectiveness analysis, 01 April 2025 – 05 April 2026.

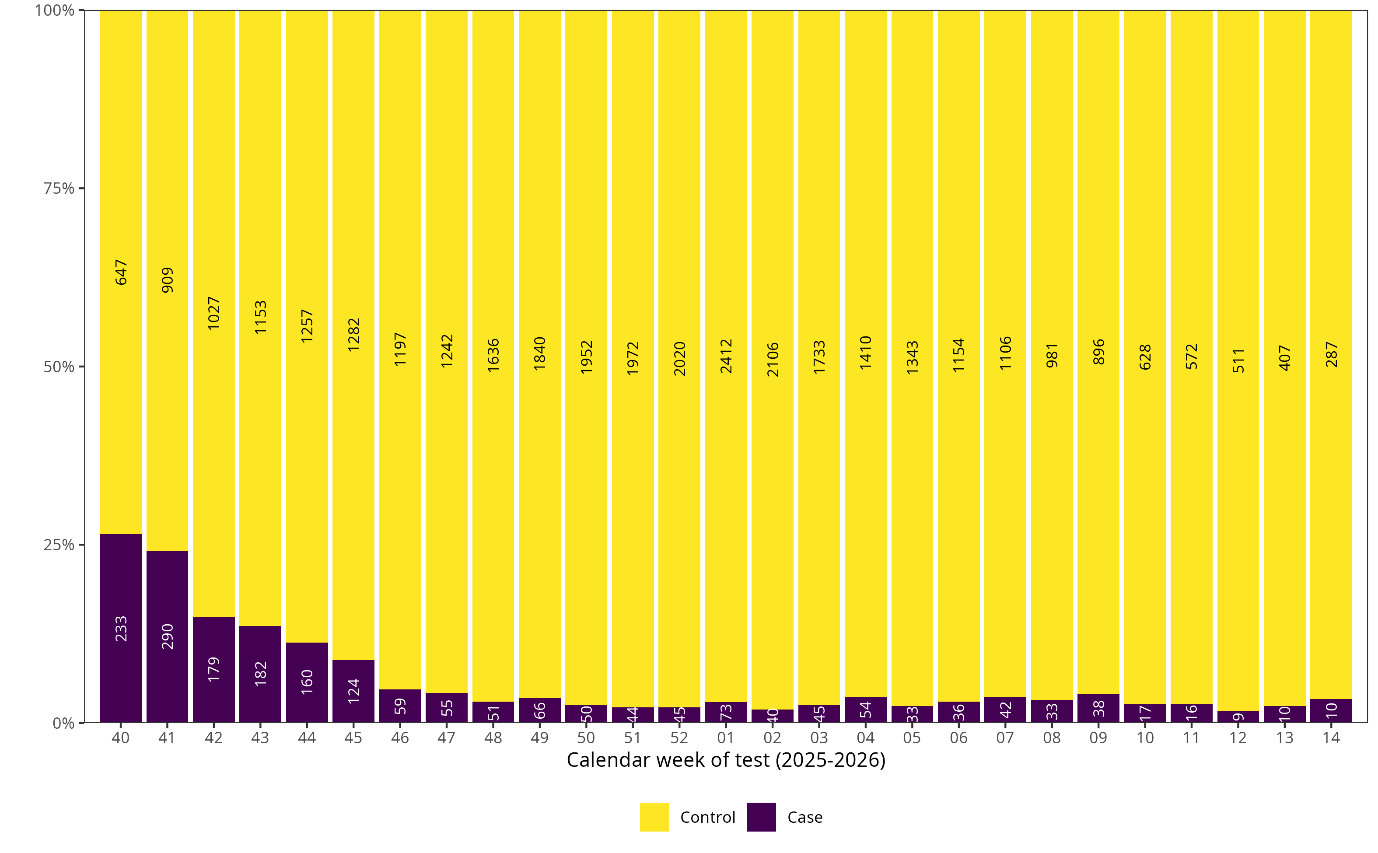

Supplementary Figure 2. Distribution of cases and controls by week of test for tests included in the autumn 2025 vaccine effectiveness analysis, 01 October 2025 – 05 April 2026.

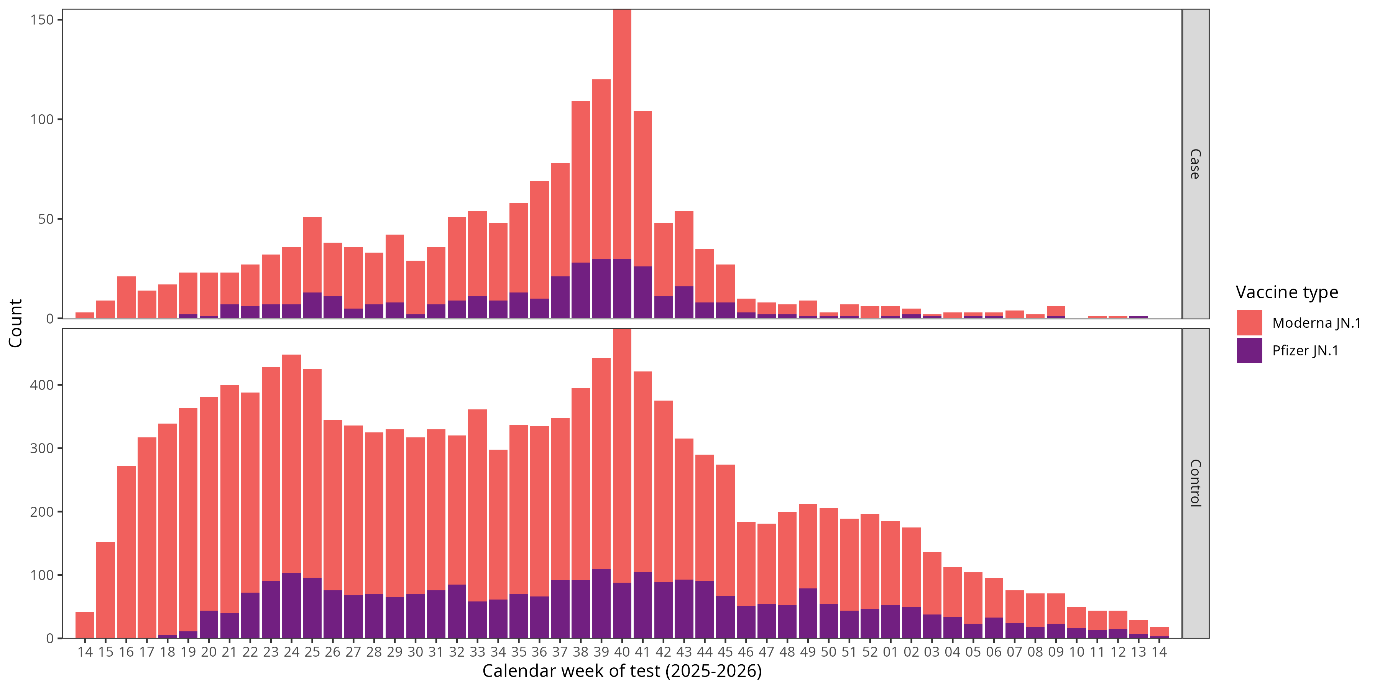

Supplementary Figure 3. Distribution of vaccinated cases and controls by week of test and vaccine manufacturer for tests included in the spring 2025 vaccine effectiveness analysis, 01 April 2025 – 05 April 2026.

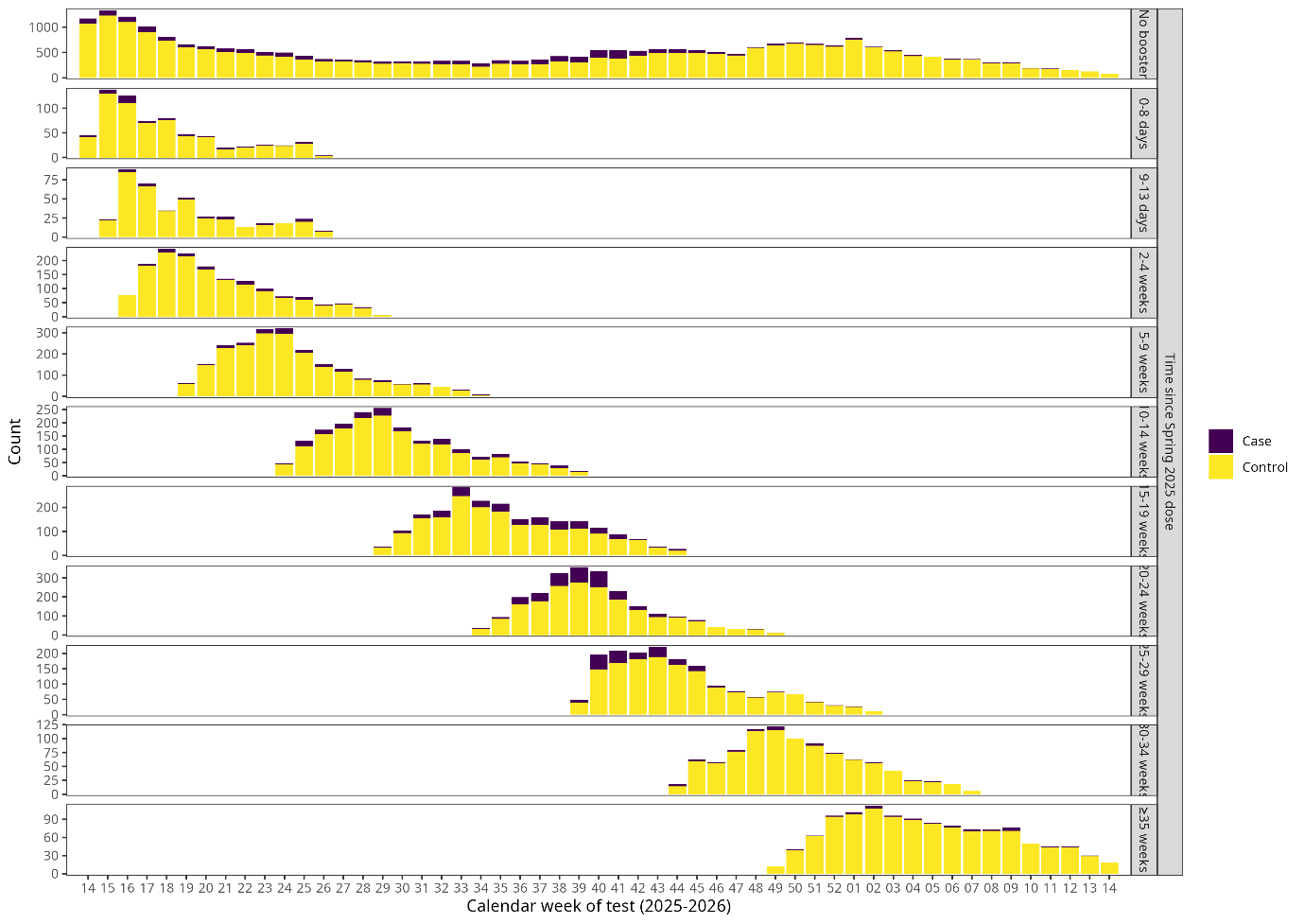

Supplementary Figure 4. Distribution of cases and controls by week of test and time since spring 2025 dose for tests included in the spring 2025 vaccine effectiveness analysis, 01 April 2025 – 05 April 2026.

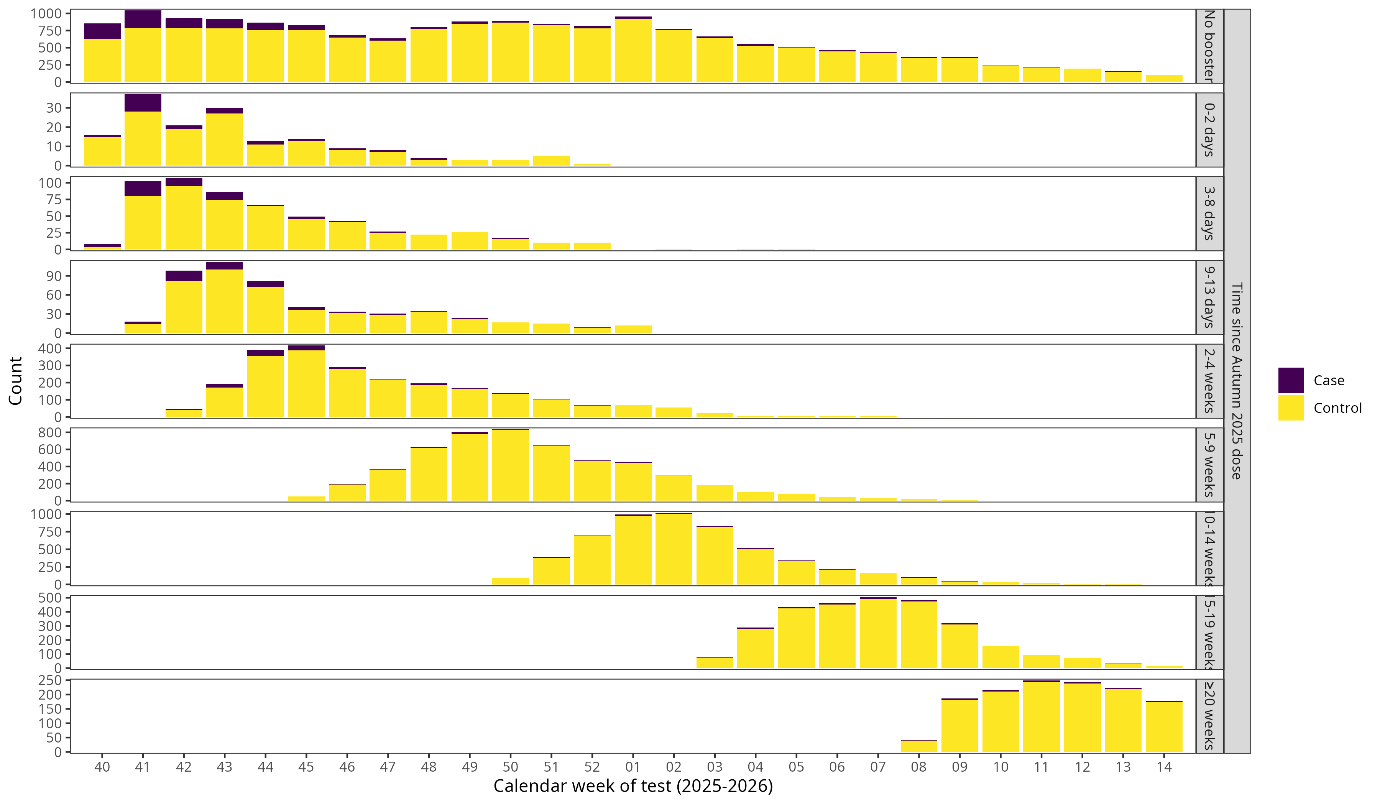

Supplementary Figure 5. Distribution of cases and controls by week of test and time since autumn 2025 dose for tests included in the autumn 2025 vaccine effectiveness analysis, 01 October 2025 – 05 April 2026.

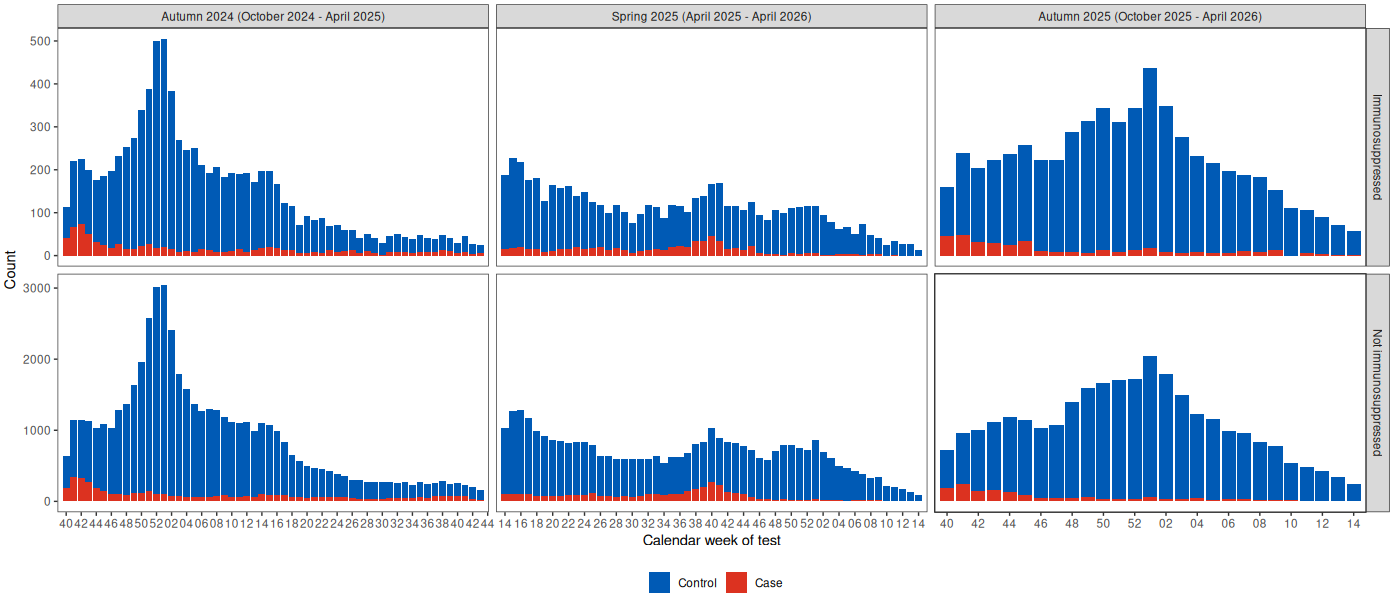

Supplementary Figure 6. Distribution of cases and controls by week of test, vaccine campaign, and immunosuppression status for tests included in the pooled vaccine effectiveness by immunosuppression status analysis, 03 October 2024 – 05 April 2026.

Supplementary Table 3. Counts of cases and controls by vaccine campaign and immunosuppression status for tests included in the pooled vaccine effectiveness by immunosuppression status analysis, 03 October 2024 – 05 April 2026.

|  | **Immunosuppressed** | | **Not immunosuppressed** | |
| --- | --- | --- | --- | --- |
| **Vaccine campaign** | Controls | Cases | Controls | Cases |
| Autumn 2024 | 7,716 | 862 | 46,122 | 4,933 |
| Spring 2025 | 5,169 | 653 | 32,256 | 3,850 |
| Autumn 2025 | 5,661 | 373 | 28,019 | 1,621 |
